# Naturally acquired immunity among Kenyan adults suppresses the West African *P. falciparum* NF54 strain in controlled human malaria infection (CHMI)

**DOI:** 10.1101/2020.08.11.20172411

**Authors:** Melissa C Kapulu, Patricia Njuguna, Mainga Hamaluba, Domtila Kimani, Joyce M. Ngoi, Janet Musembi, Omar Ngoto, Edward Otieno, Peter F Billingsley, CHMI-SIKA Study Team

## Abstract

We used controlled human malaria infection (CHMI) to study naturally acquired immunity of Kenyan adults. We administered 3.2×10^3^ cryopreserved *Plasmodium falciparum* sporozoites (SPZ, NF54 West African strain) and undertook clinical monitoring and serial quantitative PCR (qPCR). Of the 142 volunteers who were eligible for analysis: 26 (18.3%) had febrile symptoms and were treated; 30 (21.1%) reached ≥500 parasites/μl and were treated; 53 (37.3%) had parasitaemia without meeting thresholds for treatment and; 33 (23.2%) remained qPCR negative. We find that the immunity acquired by some Kenyan adults can completely suppress *in vivo* growth of a parasite strain originating from outside Kenya.

## Introduction

Humans become immune to *Plasmodium falciparum* malaria following repeated exposure^1^. Immuno-epidemiological studies show associations between immune responses and protection against malaria in the field^2^, but their interpretation is complicated by heterogeneity of exposure^3^. Furthermore, malaria is genetically diverse and vaccines that protect against heterologous parasites are required for sustained public health impact. Exposure in the field is usually with parasites of unknown genotype, further complicating inferences on protective immunity. An experimental design with controlled exposure to parasites is needed for greater confidence in inferring causality and to directly test for heterologous immunity.

Human infection studies in which investigators intentionally infect healthy volunteers have been used to understand pathogenesis, immunity, and genetic resistance to infection, and to measure the efficacy of drugs and vaccines^4^. The majority of controlled human malaria infection (CHMI) studies done to date have used *P. falciparum* (Pf)-infected mosquitoes to assess vaccine-induced efficacy in non-malaria endemic populations. The availability of aseptic, purified cryopreserved Pf sporozoites (SPZ) (PfSPZ Challenge), administered by needle and syringe has facilitated CHMI studies among non-endemic and endemic populations^5-7^. An intravenous dose of 3.2×10^3^ PfSPZ leads to an established infection with exponential *in vivo* parasite growth in 100% of volunteers from malaria non-endemic areas^8,9^.

PfSPZ Challenge has now been used in several malaria endemic populations to evaluate vaccine efficacy^10^, naturally-acquired immunity^11^, and innate resistance to infection^12^. A total of 150 volunteers in four malaria endemic countries (Gabon, Gambia, Kenya, and Tanzania), have been enrolled in published CHMI studies using PfSPZ Challenge to either assess vaccine efficacy or infectivity^6,7,10-12^. A further 300 volunteers have been or will be enrolled in vaccine efficacy CHMI trials in West and East Africa over the next 2-3 years^13^. Most of the volunteers in these CHMI studies were from urban areas (i.e. with limited prior malaria exposure). Nevertheless this limited prior exposure led to CHMI outcomes in these individuals that were different from those seen in non-endemic regions^6,7,11,12^. Ten of 43 volunteers did not develop positive blood smears for malaria parasites in studies in Tanzania^6^ and Gabon^12^, whereas 100% of volunteers in non-endemic areas develop positive blood smears. In a vaccine efficacy study conducted in Tanzania^10^, infectivity detectable by blood smear microscopy in the control group was observed in 16 of 18 volunteers. In Kenya, Nairobi, one of 28 volunteers was negative by blood smear microscopy, and showed minimal parasite growth by serial quantitative polymerase chain reaction (qPCR)^7^. In The Gambia, 17 of 19 volunteers developed infection detectable by microscopy, one was negative by microscopy but positive by qPCR, and another one was negative by qPCR^11^.

For endpoint assessment in CHMI studies, the conventional method for monitoring parasitaemia has been thick blood smear microscopy which does not detect low-density parasitaemia^14^. Molecular biomarker methods such as qPCR are substantially more sensitive^15^, and therefore provided more information on parasite growth in the CHMI studies referenced above in Kenya and in Gambia. The biomarker-based approach was recently been qualified by the USA Food and Drug Administration (FDA) as a replacement for thick blood smears in non-endemic CHMI studies^16^.

To systematically explore qPCR outcomes following CHMI in semi-immune adults, we conducted the largest CHMI study to date, recruiting volunteers with previous exposure to malaria from specific rural areas in Kenya. As adults in malaria-endemic regions of Kenya are frequently found to be asymptomatic despite significant blood parasitaemia^17^, we adopted a relatively high threshold (500 parasites per μL) for the study endpoint. This threshold is higher than that used for CHMI in non-endemic areas (where thick blood smear microscopy has been the criteria for treatment, equivalent to ~5-50 parasites per μL)^15^. However we expected parasites would be well tolerated among Kenyan adults, since the threshold is five times below the parasitaemia associated with symptomatic malaria in Kenyan children (i.e. 2,500 parasites per μL)^17^. We included criteria for treatment at lower parasite densities in the presence of significant clinical symptoms.

The aim of this study was to investigate how the *in vivo* parasite growth in CHMI would be modified by pre-existing immunity, where qPCR was used to quantify parasite growth and PfNF54 parasites were used for challenge. NF54 is a laboratory-adapted parasite line that is of West African origin and is genetically distinct from East African parasites^18^.

## Results

### Study design and study population

504 volunteers were recruited from Ahero, Kilifi North, and Kilifi South locations in Kenya and assessed for eligibility. Of these, 161 healthy volunteers were enrolled into three successive cohorts beginning in August 2016, February 2017, and February 2018 (N=37, 64 and 60, respectively) representative of volunteers from Ahero (N=15), Kilifi North (N=34), and Kilifi South (N=112). All 161 volunteers were inoculated with 3.2×10^3^ PfSPZ of PfSPZ Challenge NF54 by DVI and monitored for outcomes by clinical assessment and qPCR. All volunteers completed CHMI and were successfully treated and discharged from inpatient care (**Fig. 1**). One hundred and fifty-eight volunteers completed follow-up at day 35 post-CHMI (day 90 for the 2016 cohort). Two volunteers moved away from the study area, and one was not available for follow-up visit(s) after CHMI was completed. Data from these two volunteers were included for analysis of CHMI outcomes. Seven volunteers were found to have non-PfNF54 strain parasites (**Supplementary File 1**) and excluded from further analysis, and a further twelve volunteers were excluded on the basis of anti-malarial drug concentrations (see below) (**Supplementary File 2**). Thus, the safety analysis is reported for all 161 volunteers who underwent CHMI and qPCR outcomes are described for 142 volunteers (**Fig. 1**). The mean age at enrolment was 29.5 years old (SD 7.1) and most volunteers were male (70.2%) (**Table 1**). The baseline characteristics of the 161 enrolled volunteers are shown in **Table 1**.

**Fig. 1.**
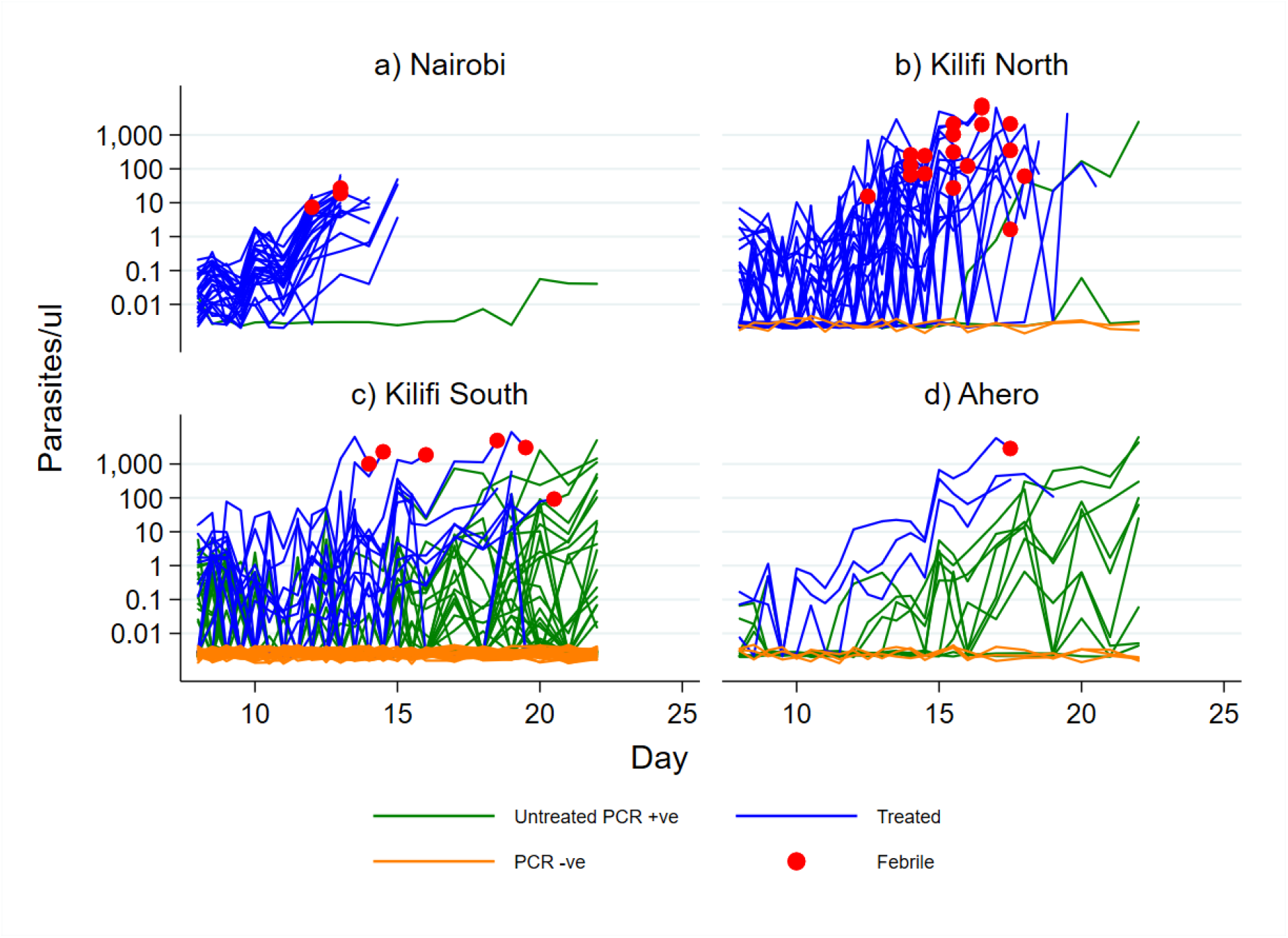
Study design and volunteer eligibility and enrolment for CHMI. ^1^One volunteer was both HIV and Hepatitis B positive; ^2^Volunteers were assessed in the fourth quarter of 2017 but were not enrolled for CHMI due to national security reasons; *Volunteers were deemed to have completed CHMI if they had received endpoint treatment. qPCR outcomes are presented for 142 after exclusions for parasite genotype and anti-malarial drug levels.

**Table 1.** Volunteer baseline characteristics based on location.

|  | **Ahero (N=15)** | **Kilifi North**<br>**(N=34)** | **Kilifi South**<br>**(N=112)** | **Total (N=161)** |
| --- | --- | --- | --- | --- |
| Age, years | 30.7 (7.1) | 28.5 (7.6) | 29.7 (6.7) | 29.5 (7.1) |
| Sex, female | 46.7% (7/15) | 20.6% (7/34) | 30.4% (34/112) | 29.9% (48/161) |
| BMI, kg/m<sup>2</sup> | 24.8 (6.9) | 22.1 (3.5) | 23.1 (3.7) | 23.1 (4.1) |
| qPCR<br>positive* | 0.0% (0/15) | 5.8% (2/34) | 25.0% (28/112) | 18.6% (30/161) |

BMI, Body mass index. qPCR, quantitative polymerase chain reaction. Data are presented as either mean and (SD) or % and (n/N). \*qPCR positive at screening and were treated with Artemether for 7 days and qPCR confirmed to have cleared parasites either at C-9 or C-3.

### Adverse events

There were no serious adverse events reported. Adverse event reporting was divided into three separate time periods: (1) the first six days from day of administration of PfSPZ (1-7), where there were few grade 1 local adverse events relating to the injection site and some isolated unsolicited events; (2) Day 8 after administration of PfSPZ up to day of treatment (8-DoT) when systemic febrile events (e.g. headache followed by fever, myalgia, and sweating) became more common; and (3) day of treatment to exit from in-patient stay (DoT-Exit) when systemic febrile adverse events were most common (**Table 2**). Unsolicited adverse events reflected symptoms of febrile malaria with a few additional events at low frequency. Solicited and unsolicited adverse events were all found to be mild (i.e. grade 1). The most common adverse event reported was headache followed by fever, myalgia, and sweating. The most commonly used concomitant medications were anti-pyretics (**Supplementary Table 2**).

**Table 2.** Adverse events.

|  |  | <sup>1</sup>C+1-C+7 | <sup>1</sup>C+8-DoT | <sup>2</sup>DoT-Exit | Total |
| --- | --- | --- | --- | --- | --- |
| Solicited Systemic | Headache | 0 | 29 (18) | 54 (33.5) | 66 (41) |
|  | Fever | 0 | 18 (11.2) | 28 (17.4) | 39 (24.2) |
|  | Fatigue | 0 | 15 (9.3) | 29 (18) | 36 (22.4) |
|  | Myalgia | 0 | 8 (5) | 15 (9.3) | 18 (11.2) |
|  | Arthralgia | 0 | 7 (4.3) | 16 (9.9) | 20 (12.4) |
|  | Chills | 0 | 7 (4.3) | 16 (9.9) | 19 (11.8) |
|  | Sweating | 0 | 7 (4.3) | 21 (13) | 25 (15.5) |
|  | Low back pain | 0 | 6 (3.7) | 10 (6.2) | 14 (8.7) |
|  | Abdominal pain | 0 | 4 (2.5) | 10 (6.2) | 13 (8.1) |
|  | Rigor | 0 | 4 (2.5) | 13 (8.1) | 14 (8.7) |
|  | Anorexia | 0 | 3 (1.9) | 9 (5.6) | 11 (6.8) |
|  | Diarrhoea | 0 | 2 (1.2) | 4 (2.5) | 6 (3.7) |
|  | Nausea | 0 | 2 (1.2) | 8 (5) | 10 (6.2) |
|  | Vomiting | 0 | 1 (0.6) | 1 (0.6) | 2 (1.2) |
| Solicited Local | Pain at the injection site | 3 (1.9) | 2 (1.2) | 0 | 5 (3.1) |
|  | Swelling | 1 (0.6) | 0 | 0 | 1 (0.6) |
|  | Redness | 2 (1.2) | 0 | 0 | 2 (1.2) |
| Unsolicited <sup>3</sup> | Headache | 7 (4.3) | 21 (13) | 41 (25.5) | 60 (37.3) |
|  | URTI | 5 (3.1) | 8 (5) | 3 (1.9) | 15 (9.3) |
|  | Others<sup>4</sup> | 2 (1.2) | 15 (9.3) | 9 (5.6) | 26 (16.1) |
|  | Abdominal pain | 1 (0.6) | 5 (3.1) | 6 (3.7) | 12 (7.5) |
|  | Arthralgia | 1 (0.6) | 7 (4.3) | 17 (10.6) | 24 (14.9) |
|  | Ear pain discomfort | 1 (0.6) | 5 (3.1) | 2 (1.2) | 7 (4.3) |
|  | Pain injection site | 1 (0.6) | 2 (1.2) | 0 | 3 (1.9) |
|  | Tinea | 1 (0.6) | 6 (3.7) | 1 (0.6) | 8 (5) |
|  | Chills | 0 | 4 (2.5) | 8 (5) | 12 (7.5) |
|  | Diarrhoea | 0 | 1 (0.6) | 1 (0.6) | 2 (1.2) |
|  | Fatigue | 0 | 10 (6.2) | 16 (9.9) | 25 (15.5) |
|  | Fever | 0 | 14 (8.7) | 19 (11.8) | 32 (19.9) |
|  | Low back pain | 0 | 4 (2.5) | 5 (3.1) | 9 (5.6) |
|  | Malaria | 0 | 1 (0.6) | 4 (2.5) | 5 (3.1) |
|  | Myalgia | 0 | 3 (1.9) | 14 (8.7) | 17 (10.6) |
|  | Nausea | 0 | 2 (1.2) | 3 (1.9) | 5 (3.1) |
|  | Rigors | 0 | 2 (1.2) | 9 (5.6) | 11 (6.8) |
|  | Sweating | 0 | 4 (2.5) | 12 (7.5) | 15 (9.3) |
|  | Swollen Limbs<sup>5</sup> | 0 | 1 (0.6) | 1 (0.6) | 2 (1.2) |
|  | Toothache | 0 | 1 (0.6) | 2 (1.2) | 3 (1.9) |
|  | Vomiting | 0 | 1 (0.6) | 0 | 1 (0.6) |

<sup>1</sup>C+1-C+7 refers to day 1 to day 7 after challenge, C+8-DoT refers to day 8 after challenge to day of treatment; <sup>2</sup>DoT-Exit refers to period of observed treatment to in-patient exit; <sup>3</sup>There are instances of double reporting of the adverse events; <sup>4</sup>Other events include tinea, dental caries, oral candidiasis, neck pain, rash, swollen left eye, hematoma at venipuncture site etc.; <sup>5</sup>One participant had an ankle sprain possibly due to sports during inpatient stay. \*Percentages expressed over the total sample challenged (N=161); All events reported were grade 1; Median duration of AE's in days: 1; min duration of AE's in days: <1; max duration of AE's in days: 70days but when excluding Tinea (11days).

The most common abnormalities on safety blood tests during CHMI were Grade 1 and 2 elevated alanine aminotransferase (ALT) and creatinine values, all of which resolved to within the normal range during the follow-up period (**Supplementary Table 3**). ALT increases were more common in males compared with females (**Supplementary Fig. 1**). Markedly reduced platelet counts (Grade 4) were observed in two volunteers, as has previously been described in malaria^19^. There were no signs of any bleeding tendency, consistent with previous reports of severe thrombocytopenia in malaria^19^.

### Assessment of presence of anti-malarial drugs

Anti-malarial drugs were detected in some volunteers (**Table 3** and **Supplementary Fig. 3**). Two volunteers had detectable concentrations of chloroquine (i.e. 3.6 ng/ml and 5 ng/ml) prior to CHMI, both at concentrations well below the reported minimum inhibitory concentration (MIC) of 67 ng/ml^20^ (i.e. at 5 ng/ml plasma and 3.6 ng/ml plasma); one of these met criteria for treatment before Day 22, the other did not. Twelve volunteers had lumefantrine concentrations above the reported MIC of 280 ng/ml plasma^21^, none of whom met criteria for treatment. A further 64 volunteers had concentrations below the MIC, of whom 43 met criteria for treatment before day 22, and 21 did not. Thus, among volunteers with detectable lumefantrine concentrations (albeit below the MIC), criteria for treatment were met slightly less often than among volunteers in whom lumefantrine was undetectable, 33.4% (25/73) vs 43.5% (33/76) (p=0.25, **Table 3**).

**Table 3:** Measured anti-malarial drug concentration by CHMI outcome.

|  | MIC (ng/ml) | Volunteers meeting treatment criteria with >MIC | Volunteers meeting treatment criteria with <MIC | Volunteers meeting treatment criteria with undetectable levels | *p* value (<MIC vs undetectable) |
| --- | --- | --- | --- | --- | --- |
| Artesunate | N/A | 0.0% (0/0) | 0.0% (0/0) | 36.1% (58/161) |  |
| Chloroquine | 67 | 0.0% (0/0) | 50.0% (1/2) | 35.9% (57/159) | 0.7 |
| Lumefantrine | 200 | 0.0% (0/12) | 33.4% (25/73) | 43.5% (33/76) | 0.25 |
| Pyrimethamine | 100 | 0.0% (0/1) | 0.0% (0/1) | 36.3% (58/160) | 0.45 |
| Sulfadoxine | 100 | 66.7% (2/3) | 33.4% (1/3) | 35.5% (56/158) | 0.26 |

MIC: minimum inhibitory concentration based on published literature; N/A not applicable. Data are presented as % and (n/N) or otherwise specified. MICs are not well defined for Pyrimethamine or Sulfadoxine, and an arbitrary value is chosen based on levels seen in the first 24 hrs after sulfadoxine pyrimethamine use in adult volunteers.

One volunteer had high concentrations of both pyrimethamine and sulfadoxine and did not meet criteria for treatment. Two further volunteers had detectable concentrations of sulfadoxine without pyrimethamine and both met criteria for treatment before Day 22. There was no detectable artemether or dihydroartemisinin in any sample. The prevalence of detectable lumefantrine concentrations was higher in Kilifi South (62.5%, 70/112) compared to Kilifi North (20.6%, 7/34). Lumefantrine concentrations measured at the two independent laboratories were closely correlated (r=0.93, *p*<0.0001).

### Analysis of qPCR outcome following CHMI

For further analysis of qPCR outcomes by location we therefore excluded volunteers with lumefantrine concentrations above the MIC and we excluded the volunteer with high pyrimethamine and sulfadoxine concentrations. We did not find concentrations of chloroquine or of lumefantrine below the MIC to be strongly associated with outcome and therefore retained data from volunteers with these drug levels, therefore and retained volunteers with undetectable drug levels. Thus, we included data from 142 volunteers for further analysis (**Table 4**). One volunteer requested early treatment at C+21 therefore was missing qPCR data from C+22. The volunteer was qPCR negative up to day 21, and it was therefore assumed they would be negative at day 22 and their data were included in the final analysis.

**Table 4:** Malaria diagnosis outcome based on location.

|  | Nairobi (N=28) | Ahero (N=15) | Kilifi North (N=34) | Kilifi South (N=93) |
| --- | --- | --- | --- | --- |
| PCR (-) | 0.0% (0/28) | 20.0% (3/15) | 5.9% (2/34) | 30.1% (28/93) |
| Treated, febrile | 11.1% (3/28) | 6.7% (1/15) | 52.9% (18/34) | 6.5% (6/93) |
| Treated, non-febrile | 85.7% (24/28) | 13.3% (2/15) | 35.3% (12/34) | 11.8% (11/93) |
| Untreated but PCR (+) | 3.5% (1/28) | 60.0% (9/15) | 5.9% (2/34) | 51.6% (48/93) |

Data are presented as % and (n/N) or otherwise specified. PCR (-) refers to PCR negative; PCR (+) refers to PCR positive.

We combined our data from Kilifi and Ahero with data from a previous pilot CHMI study conducted in Nairobi (an urban area with no malaria transmission)^7^. The qPCR results over time in relation to location, outcome, and febrile status are shown in **Fig. 2**. Heterogeneous patterns of parasite growth were observed over time with: (a) some volunteers showing rapid and consistent growth; (b) others showing early parasite growth that subsequently appeared to be suppressed; (c) late growth that was inconsistent and did not reach the threshold criteria for treatment; or (d) negative qPCR results throughout the period of monitoring. Individual qPCR results for each volunteer are shown as supplementary figures for the respective locations - Nairobi, Ahero, Kilifi North, and Kilifi South, (**Supplementary Figure 3**). In addition, individual qPCR results are shown for each respective successive cohort 2016, 2017 and 2018 (**Supplementary Figure 4**).

**Fig. 2.**
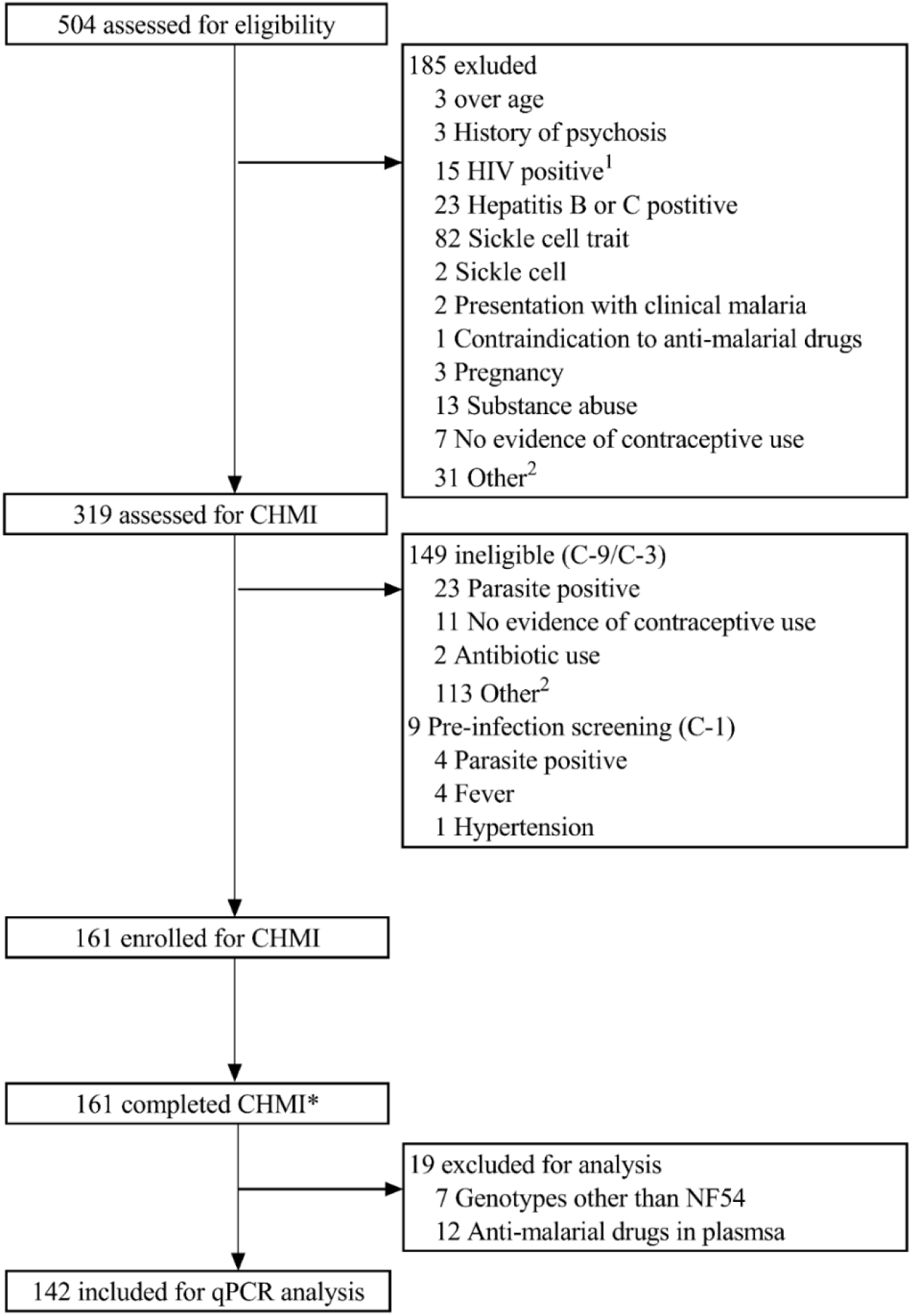
qPCR outcome based on volunteer location. Blood samples from C+8 (C+7.5 for Nairobi) onwards after inoculation to determine parasitaemia from: (A) Nairobi – N=27; (B) Kilifi North – N=34; (C) Kilifi South – N=93; and (D) Ahero – N=15. Parasitaemia was determined by asexual *18S* ribosomal RNA gene qPCR done in Kilifi. Blue lines represent individuals who required treatment and reached treatment threshold (reached DoT); green lines represent individuals who did not meet criteria for treatment threshold but were qPCR positive; orange lines represent individuals who were qPCR negative throughout monitoring; and red dot denotes individuals were febrile and met treatment criteria.

Of the 142 volunteers resident in Kilifi or Ahero (i.e. excluding Nairobi residents): 26 (18.3%) had febrile symptoms and were treated; 30 (21.1%) reached ≥500 parasites/μl and were treated; 53 (37.3%) had parasitaemia without meeting thresholds for treatment and; 33 (23.2%) remained qPCR negative, 10 of these with no detectable antimalarial drug levels (**Supplementary Table 4**). Of the 28 volunteers previously included in CHMI in Nairobi, all experienced parasite growth with 27 (96.4%) meeting the criteria for malaria diagnosis and one volunteer (3.6%) had parasitaemia without meeting thresholds for treatment.^7^ **Table 5** shows the distribution of these qPCR outcomes by location. Criteria for treatment were more frequently met among volunteers from Nairobi and Kilifi North, and less frequently met for volunteers from Ahero or Kilifi South (**Table 5**).

In contrast, in the high transmission locations of Kilifi South and Ahero it was more common to observe volunteers that experienced limited parasite growth or with negative qPCR results. The presence of fever was more common in Kilifi North than in Kilifi South or in Ahero. Fever was less frequent in the Nairobi study where treatment was given based on microscopy (microscopy is positive at 50 parasites/μl by qPCR assay^14^ but volunteers were treated at 500 parasites/μl blood in the present study). Similar results were seen after excluding all volunteers with detectable lumefantrine concentrations (**Supplementary Table 4**).

### PCR parasite detection comparisons

A subset of samples from the 2018 enrolment cohort, a total of 120 samples corresponding to 20 volunteers from time points C+8 to C+10.5 (8 volunteers who were qPCR negative throughout in Kilifi and 12 other volunteers with positive readings), were shared with two external laboratories at Mahidol University (MU), Thailand and at University of Washington (UW), USA for their analytically sensitive qPCR and reverse transcription PCR (RT-PCR) assays, respectively^15,16,22^. There was close agreement of results from the Kilifi laboratory assay with those of MU (r=0.65, *p*<0.0001) and UW (r=0.64, *p*<0.0001), and between MU and UW assays (r=0.79, *p*<0.0001) (**Supplementary Figure 5**). Of the eight volunteers considered qPCR negative in the Kilifi assay, five had parasitaemia detected at low levels in the MU assay and six had low-level parasitaemia in the UW assay (**Supplementary Figure 6**). Two of the eight volunteers were negative for all the three assays whilst one volunteer was positive for the UW assay but negative for the other two assays.

Finally, we re-analysed the ALT elevations observed after CHMI according to qPCR outcomes and found that ALT elevations did not vary according to qPCR status. Volunteers who were qPCR negative for malaria parasites following CHMI had a median ALT of 46 (IQR 27-120) at Day 10, compared with a median ALT of 43 (IQR 26-80) among those who were qPCR positive (Z=0.79, *p*=0.43) (**Supplementary Figure 7**).

## Discussion

We found CHMI to be safe and well tolerated in the 161 volunteers enrolled in this study who were recruited from malaria-endemic regions of Kenya. Using qPCR, we show that outcomes following CHMI vary depending on previous history of malaria exposure. In malaria non-endemic locations, inoculation with the same dose of 3.2×10^3^ of PfSPZ Challenge by DVI that was used in our study leads to infection and *in vivo* parasite growth in 100% of volunteers^8,9^. In our study with Kenyan volunteers, we observed *in vivo* parasite growth leading to parasite densities meeting treatment criteria in only 39.5% of our volunteers (i.e. 18.3% with symptoms plus 21.1% without). In 37.3% of the volunteers we observed limited *in vivo* growth that was partially suppressed such that treatment criteria were not met. In another 23.3% of volunteers we observed complete suppression of parasite growth such that parasites were not detected by our qPCR assay. The challenge strain is genetically distant to East African parasites, implying that the complete suppression of parasite growth was the result of heterologous immunity.

This study provides the largest dataset to date on CHMI outcomes in an adult population from a malaria-endemic area. The CHMI outcomes in participants from different locations clearly followed the underlying intensity of malaria exposure (i.e. with Nairobi appearing similar to non-endemic countries, and then Kilifi North, Kilifi South, and Ahero associated with increasing control of parasite growth). It is likely that prior malaria exposure rather than human genetic differences explain the observed variations in CHMI outcomes.

In this study, anti-malarial drugs were detectable in a significant proportion of volunteers, particularly lumefantrine, which has a relatively long half-life^23^. Thus, we excluded volunteers with baseline concentrations of lumefantrine above the reported minimum inhibitory concentration (MIC) for *P. falciparum^21^*. There was no evidence that drug concentrations explained outcomes among the subjects with lumefantrine concentrations below the MIC, and they were therefore retained in the analysis. Furthermore, there was a similar pattern of results seen when we re-analyzed data after excluding all volunteers with detectable lumefantrine concentrations (**Supplementary Table 4**). Artemether-lumefantrine is widely available in Kenya, and is the likely source of lumefantrine in our volunteers^24^. Artemisinins were not detected in any volunteers, and these drugs have a very short half-life^25^. The observed concentrations of lumefantrine were low or undetectable. Taking this together with the absent artemisinin plasma concentrations, we conclude that any putative doses of artemether-lumefantrine were likely taken several weeks prior to CHMI. Although we solicited for history of anti-malarial use during the screening visit and excluded volunteers who reported recent treatment, volunteers may have previously taken medication with little or no explanation from medical staff. We therefore believe our findings are due to inadvertent or forgotten previous use of anti-malarial drugs rather than recent surreptitious anti-malarial use to influence outcomes of CHMI.

In some volunteers Kilifi qPCR results were negative throughout. The Kilifi assay has an analytical sensitivity of 20 estimated parasites/ml. Using a 0.5 or 1 ml sample, the UW qRT-PCR and the MU qPCR assays are both more sensitive than the Kilifi qPCR to parasite densities below 20 parasites/ml^15,22^. Using these more sensitive assays, 6 of 8 volunteers who were considered qPCR negative in Kilifi were found to have low level parasitemia by UW or MU assays. If we had assessed all 33 Kilifi qPCR volunteers with UW and MU assays, we might expect the negative qPCR rate to be reduced by a similar proportion from 23% to 6%. Repeated assays of increased blood sample volumes would likely further increase the sensitivity for detecting low density parasitaemia^15,22^. However, since two of eight volunteers’ samples that negative with all three qPCR assays, we cannot exclude the possibility of sterile pre-erythrocytic immunity in some volunteers. Furthermore, that parasites were undetectable in some volunteers immediately after emergence from the liver (i.e. 7.5-9.5 days post-CHMI) and then subsequently detectable suggests a low liver-to-blood inoculum^26^, which would indicate a partial role for pre-erythrocytic-stage immune responses, albeit not necessarily sterile protection. Nevertheless, the suppression of parasite growth by host immunity is profound.

In conclusion, we have shown CHMI studies to be safe in a malaria endemic location. Past exposure to malaria in an adult population leads to a range of outcomes in CHMI studies in malaria-endemic regions. *P. falciparum* shows marked antigenic diversity, which results in parasites evading host immunity^27^. Our data suggest that sufficiently cross-reactive blood-stage immunity can be acquired on exposure to East African parasites to clear qPCR-detectable parasitaemia from a phylogenetically distant West African parasite (i.e. NF54).

## Methods

### Study design and volunteer population

The study was open, unblinded, and non-randomised. The protocol has been published previously^13^. All volunteers received an intravenous injection (direct venous inoculation (DVI)) of 3.2×10^3^ P*f*SPZ of the West African NF54 strain (P*f*SPZ Challenge, i.e. aseptic, purified, cryopreserved P*f*SPZ). Volunteers were recruited from differing malaria endemic regions in Kenya: Ahero in Western Kenya (moderate to high transmission at community age-adjusted parasite rates (P*f*PR) of 40%); Kilifi South (moderate transmission, currently at a PfPR of 20% but historically a P*f*PR of 40%); and Kilifi North on the Kenyan Coast (low to no malaria transmission at present, but historically a PfPR of 25%)^28^. We included data from a previous study based in Nairobi (Supplementary Methods) where there is no malaria transmission^7^, although volunteers may have been exposed during previous residence or childhood elsewhere in Kenya.

The study was conducted at the KEMRI Wellcome Trust Research Programme in Kilifi, Kenya and received ethical approval from the KEMRI Scientific and Ethics Review Unit (KEMRI//SERU/CGMR-C/029/3190) and the University of Oxford Tropical Research Ethics Committee (OxTREC 2-16). The study was registered on ClinicalTrials.gov (NCT02739763), conducted based on good clinical practice (GCP), and under the principles of the Declaration of Helsinki.

### Study enrolment and administration of PfSPZ Challenge

For the current study in Kilifi, following recruitment and informed consent procedures, volunteers went through a screening process to determine their health status and past exposure to malaria. Furthermore, anti-schizont antibodies, as previously described^7^, were measured at screening to ensure that the enrolled volunteers would include a range of anti-schizont antibody titres. This was done to ensure equal enrollment from each of the three tertiles of anti-schizont antibody titres identified at screening. A dose of 3.2×10^3^ PfSPZ was administered by direct venous inoculation in 0.5 mL through a 25-gauge needle over several seconds, after which volunteers were monitored for blood parasitaemia by qPCR to determine parasite growth. Volunteers were enrolled in three successive cohorts in 2016, 2017 and 2018.

### Safety monitoring

All volunteers were monitored for any adverse events (AEs), solicited and unsolicited, for the duration of CHMI. Signs and/or symptoms of malaria were assessed and recorded. Abnormal laboratory findings were graded for severity based on population sex-specific adapted reference ranges (**Supplementary Table 1**). In keeping with the study data tabulation model implementation guide for human clinical trials, the day of injection/inoculation was defined as day 1 (C1) and not day 0 (C0).

### Quantitative PCR for parasite detection

In the Kilifi study real-time qPCR results were used for endpoint criteria. Endpoint criteria were considered met and anti-malarial treatment given when: (a) parasitaemia reached 500 parasites/μl; (b) clinically significant signs and/or symptoms were observed, with any evidence of malaria parasites by blood film positivity; or (c) at day 22 (21 days post-infection). For criteria (b) thick and thin blood films were prepared when requested by the assessing clinicians and made from whole blood by experienced microscopists who examined 100 high power fields. In Nairobi endpoint criteria were met when thick-blood film microscopy was positive with retrospective analysis of qPCR results. A positive thick film corresponds to a qPCR quantification of ~50 parasites per μL^14^.

For parasite detection, a sensitive high-volume qPCR assay, detecting the *18S* ribosomal RNA *P. falciparum* gene, was used in real time where 500 μl of whole venous blood was collected twice every day from days 8 to 15 post infection and then once every day from days 16 to 22 post-infection^13^. Details of the qPCR methods are described in the Supplementary Methods. In addition, MSP-2 genotyping and anti-malarial drug detection was carried out (Supplementary Methods).

## Data Availability

Volunteer data will be made available including data dictionaries after de-identification of the volunteer data that represent the results reported in this article. The data will be available to researchers who will need to submit proposals to to gain access to the data following a signed data access agreement. The study protocol, informed consent forms, and all other associated documents have been previously published.

## Acknowledgments

This work was supported by a Wellcome Trust grant (107499). We would like to thank all the study volunteers who participated in the CHMI-SIKA study. We are also very thankful to the larger study teams in Kilifi and Ahero specifically all the fieldworkers, health community workers who recruited volunteers, the data entry clerks, clinical, pharmacy, and laboratory teams; and the collaborating manufacturing, quality systems, regulatory, pharmaceutical operations and clinical teams at Sanaria Inc. without whom this work would not have been possible. We would also like to thank the diagnostic teams at Mahidol University and the University of Washington for their assistance. All the members of the CHMI-SIKA study team read and reviewed the manuscript. This manuscript is published with permission and/or approval of the Director KEMRI.

## Author Contributions

MCK: conceptualization, study design, data collection, data analysis, data interpretation, methodology, supervision, figures, validation, writing - original draft preparation, writing - review and editing; PN: clinical support, investigation, methodology, supervision, validation, writing - review and editing; MH: clinical support, investigation, supervision, validation, writing - review and editing; DK and JMN: data collection, investigation, methodology, validation, writing - review & editing; JM and ON: clinical support, investigation, validation, writing - review and editing; EO: data curation, data analysis, validation, figures, writing - review and editing; and PFB: supervision, validation, resources, writing - review and editing; *CHMI-SIKA Study Team Author Contributions*

AIA, PCC, KPC, ZL, SHH, IJ, DoK, SiK, RK, SaK, CK, NK, MI, JoM, VM, KSM, MoM, SCM, JeM, MiM, JdM, IN, MN, DO, DwO, JO, MOO, AMS, JS, NS, BS, JoT, JaT, JW, TNW, MW: investigation, validation, writing - review and editing; AA, FrO, and JaO: clinical support, investigation, validation, writing - review and editing; NK, KeM, DM, GN, MO: data curation, validation, writing - review and editing; YA, EJ, TLR, BKLS: supervision, validation, resources, writing - review and editing; PCB, KvM, SLH, FN, BO, and FaO: conceptualization, funding acquisition, supervision, writing - review and editing; and PB: conceptualization, study design, oversight of work, data analysis, data interpretation, funding acquisition (lead applicant for funding), figures, writing – review and editing

